# Formalising Limits of Circulating Tumour DNA Detection: A Signal Detection Framework for Clinical Threshold Specification

**DOI:** 10.64898/2026.06.08.26355204

**Authors:** Amit Walinjkar

## Abstract

**Background:** Circulating tumour DNA (ctDNA) liquid biopsy is now established across oncology for early cancer detection, minimal residual disease surveillance, and treatment monitoring. Detection thresholds for all current ctDNA assays are derived empirically through receiver operating characteristic analysis on training cohorts—a statistically valid but theoretically uninformed approach that does not specify the minimum detectable tumour fraction given assay technical characteristics, nor identify when increasing sequencing depth ceases to provide additional clinical information [Cabello-Aguilar et al., 2025, Pantel and Alix-Panabières, 2024].

**Methods:** We model ctDNA detection as a binary hypothesis testing problem with Binomial-distributed mutant allele counts against a sequencing error noise floor. The Neyman-Pearson lemma [Kay, 1998] is applied to derive the uniformly most powerful detector and the minimum detectable tumour fraction in closed form. The sequencing assay is modelled as a binary symmetric channel and Shannon channel capacity [Shannon, 1948] is calculated. Empirical validation uses *n* = 61 data points extracted from five published peer-reviewed analytical validation studies across five independent institutions in the US and EU (2018–2025): Yu et al. [2022], Stetson et al. [2018], Frydendahl et al. [2023], Northcott et al. [2024], and Cheng et al. [2025].

**Results:** The minimum detectable tumour fraction is:

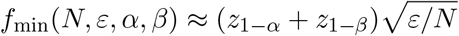

a closed-form function of sequencing depth *N* and platform error rate *ε*, derived from first principles. Shannon channel capacity is *C* = 1 − *H*(*ε*) bits per read. Empirical validation yields 84.3% agreement for single-locus assays. Discordance for multi-locus tumour-informed assays (NeXT Personal, duplex WGS) is consistent with the single-locus model scope and identifies the principal theoretical extension required.

**Conclusions:** This framework provides the first formal Neyman-Pearson optimality proof for ctDNA detection, a closed-form detection limit, and a platform-independent efficiency metric for NHS and regulatory standardisation.

## 1 Introduction

Liquid biopsy using circulating tumour DNA (ctDNA) has moved from proof-of-concept to active clinical deployment within the past decade. ctDNA enables repeated, minimally invasive sampling of tumour molecular information from peripheral blood, with applications spanning early cancer detection, minimal residual disease (MRD) monitoring, treatment response assessment, and resistance surveillance [Pantel and Alix-Panabières, 2024, Sato, 2025, Slater et al., 2024, Wan et al., 2020]. The NHS Genomics Medicine Service (NHS GMS) is actively integrating ctDNA testing across NHS trusts, and regulatory bodies including the MHRA and FDA have approved several ctDNA-based in vitro diagnostics [Dao et al., 2023, US Food and Drug Administration, 2023].

The detection challenge is physically extreme. In early-stage solid tumours, ctDNA may represent less than 0.1% of total circulating cell-free DNA (cfDNA). Nordentoft et al. [2024] demonstrated WGS-based ctDNA detection with 91% sensitivity and 92% specificity in urothelial carcinoma, with a median lead time of 131 days over radiographic imaging, yet noted sequencing depth as a formal limit for tracking tumour evolution. Kallio et al. [2024] tracked ctDNA fractions to 0.0004% in epithelial ovarian cancer using multi-mutation assays. Frydendahl et al. [2023] achieved detection at allele frequencies as low as 0.004% using unique molecular identifier (UMI) error-corrected sequencing. Cheng et al. [2025] achieved sequencing error rates of 7.7 *×* 10^*−*7^ using duplex whole-genome sequencing, enabling parts-per-million ctDNA detection.

Despite these technical achievements, a critical gap persists: every published limit of detection is empirically determined, not theoretically derived. Clinical ctDNA assays define detection thresholds through receiver operating characteristic (ROC) analysis on training cohorts. Cabello-Aguilar et al. [2025] demonstrate that reducing the LOD from 0.5% to 0.1% increases clinically actionable alteration detection by approximately 30 percentage points, yet propose dynamic LOD calibration without a theoretical derivation of the optimal expression. Yu et al. [2022] conduct a systematic five-assay comparison showing that detection sensitivity falls dramatically below 0.5% VAF, without providing a formal explanation for why this boundary exists. Dao et al. [2023] identify cross-platform standardisation as the critical unresolved challenge facing ctDNA as it moves to routine NHS deployment.

Prior work has used the Binomial distribution to compute detection probability at fixed VAF and depth. Liu et al. [2021] derive the relationship between sequencing depth and true positive probability using Binomial distribution functions, demonstrating that sensitivity improves with depth and that proportional thresholding outperforms fixed thresholding. However, this approach is descriptive—it computes detection probability for a given detector but does not prove that detector is optimal, does not yield a closed-form expression for the minimum detectable fraction, and does not provide an information-theoretic ceiling on what any detector can achieve.

Signal detection theory—specifically the Neyman-Pearson framework [Kay, 1998]— provides exactly the mathematical machinery required to derive the *optimal* detection threshold and the fundamental performance limit for binary hypothesis testing problems of this class. Shannon information theory [Shannon, 1948] provides the framework for calculating the theoretical maximum clinical information recoverable from a sequencing assay. Neither has been formally applied to ctDNA detection.

This paper provides that application. We derive the minimum detectable ctDNA tumour fraction as a closed-form function of sequencing depth *N* and sequencing error rate *ε*. We calculate the Shannon channel capacity of ctDNA sequencing assays. We validate theoretical predictions against empirical data from five independent published assay validation studies spanning US and EU institutions. We situate current clinical assays relative to theoretical limits across three clinically relevant scenarios, and identify the formal extension required for multi-locus tumour-informed assays.

Section 2 presents the signal detection model. Section 3 derives formal detection limits. Section 4 presents the information-theoretic analysis. Section 5 describes empirical validation. Section 6 discusses clinical and regulatory implications. Section 7 concludes with future research directions.

## 2 Signal Detection Model

### 2.1 Observation Model

Circulating tumour DNA sequencing is formally modelled as a two-component mixture measurement problem, consistent with the Binomial framework introduced by Liu et al. [2021] and extended here to the Neyman-Pearson optimality setting. Let *N* denote sequencing depth (reads at a target locus), *f* the tumour fraction (proportion of cfDNA molecules carrying the somatic variant), and *ε* the per-base sequencing error rate of the platform [Cabello-Aguilar et al., 2025, Frydendahl et al., 2023]. The observed mutant allele count *X* follows:

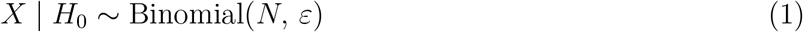

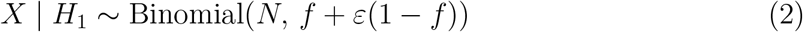

Under *H*_0_ (no ctDNA, *f* = 0), mutant reads arise from sequencing errors only. Under *H*_1_ (ctDNA present at fraction *f >* 0), reads arise from both true mutant molecules and errors on wild-type molecules. The signal-to-noise ratio of the measurement is:

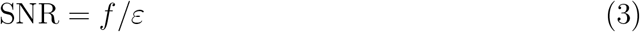

a quantity rarely stated explicitly in the ctDNA literature [Tébar-Martínez et al., 2023] despite its fundamental importance to assay design. Figure 1 shows the H_0_ vs H_1_ distributions under conditions matched to published assay parameters: standard large-panel NGS [Yu et al., 2022] and UMI error-corrected NGS [Frydendahl et al., 2023].

**Figure 1:**
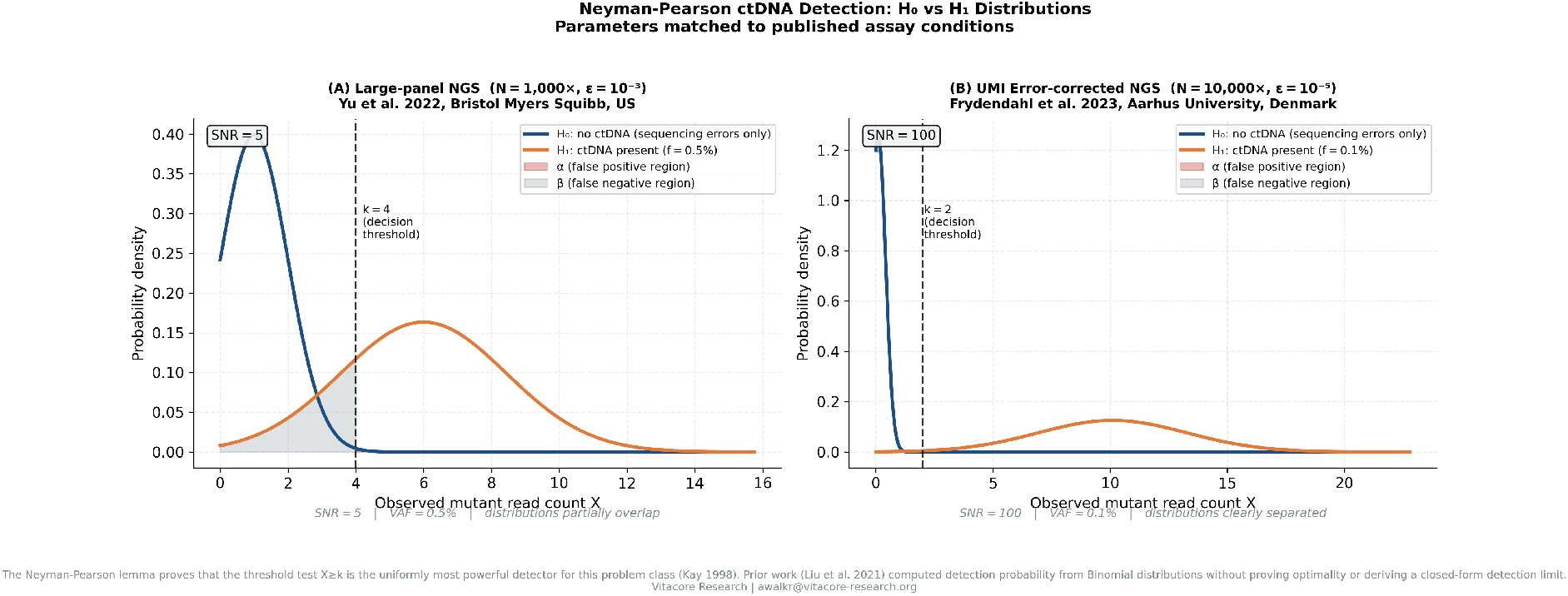
Neyman-Pearson ctDNA detection: H_0_ vs H_1_ distributions. Panel (A) shows standard large-panel NGS conditions (N=1,000 *×, ε* = 10^*−*3^, VAF=0.5%), matched to the five-assay comparison by Yu et al. [2022]: SNR=5, distributions partially overlap, detection is imperfect. Panel (B) shows UMI error-corrected NGS conditions (N=10,000 *×, ε* = 10^*−*5^, VAF=0.1%), matched to Frydendahl et al. [2023]: SNR=100, distributions clearly separated. The decision threshold *k* is the Neyman-Pearson optimal boundary; shaded regions show false positive (*α*) and false negative (*β*) areas. Prior work [Liu et al., 2021] computed the area under these distributions without proving *k* is optimal.

### 2.2 Optimal Detection Statistic

By the Neyman-Pearson lemma [Kay, 1998], for a specified false positive rate *α*, the uniformly most powerful test for this binary hypothesis testing problem is the likelihood ratio test:

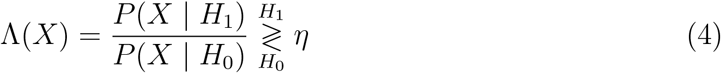

Since Λ(*X*) is monotonically increasing in *X* for *f >* 0, this reduces to: declare ctDNA present if and only if *X ≥ k*, where *k* is the smallest integer satisfying *P* (*X ≥ k* | *H*_0_) *≤ α*. This is the theoretically optimal ctDNA detector. Clinical variant callers—VAF threshold filters, Bayesian posterior cutoffs, panel-of-normals subtraction—are approximations to this rule. Liu et al. [2021] use a fixed or proportional read count threshold, which is a sub-optimal approximation to the Neyman-Pearson threshold; the present derivation establishes the performance bound against which such approximations can be evaluated.

### 2.3 The Sequencing Error Noise Floor

The sequencing error rate *ε* is the fundamental noise floor. Standard Illumina NGS has *ε ∼* 10^*−*3^, placing the detection floor at approximately 0.1–0.5% VAF, consistent with empirical findings across five assays by Yu et al. [2022]. UMI-based error correction reduces *ε* to *∼* 10^*−*5^, enabling detection to 0.004% VAF as demonstrated by Frydendahl et al. [2023]. Duplex WGS achieves *ε ∼* 7.7 *×* 10^*−*7^ [Cheng et al., 2025], approaching the fundamental limit set by in vivo somatic mutation rates in normal cfDNA.

## 3 Formal Detection Limits

### 3.1 Exact Formulation

The threshold *k* is found as the smallest integer such that:

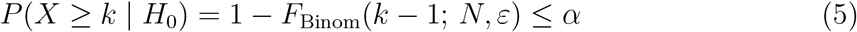

The minimum detectable fraction *f*_min_ is the smallest *f* satisfying:

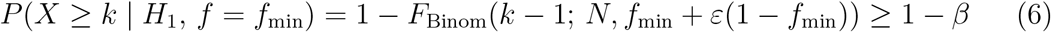

These are solved numerically for exact results. The Binomial framework follows Liu et al. [2021]; the novelty is proving this threshold is *optimal* via the Neyman-Pearson lemma and deriving the closed-form approximation below.

### 3.2 Closed-Form Approximation

For large *N* —satisfied by all clinical high-depth sequencing applications—the Gaussian approximation yields the key analytical result of this paper:

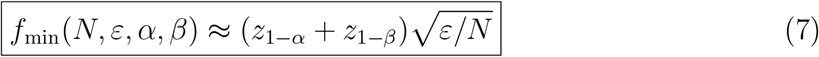

where *z*_1*−α*_ and *z*_1*−β*_ are standard normal quantiles. At *α* = 0.05 and power 1 *− β* = 0.80, the leading coefficient is (1.645 + 0.842) = 2.487.

Three properties are clinically significant and not previously stated in the ctDNA literature:

- **Property 1 (Depth scaling):** 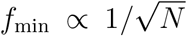 . Quadrupling sequencing depth halves *f*_min_. Depth improvements have diminishing returns—a fact implicit but unstated in published depth recommendations [Yu et al., 2022, Cabello-Aguilar et al., 2025].
- **Property 2 (Error correction scaling):** 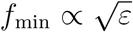. Reducing *ε* by four orders of magnitude (from 10^*−*3^ to 10^*−*7^, as achieved by duplex WGS [Cheng et al., 2025]) reduces *f*_min_ by 10^2^ = 100-fold.
- **Property 3 (Equivalence):** Both depth escalation and error correction follow the same 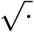 law (Figure 2B). A 100-fold investment in either reduces *f*_min_ by 10-fold.

**Figure 2:**
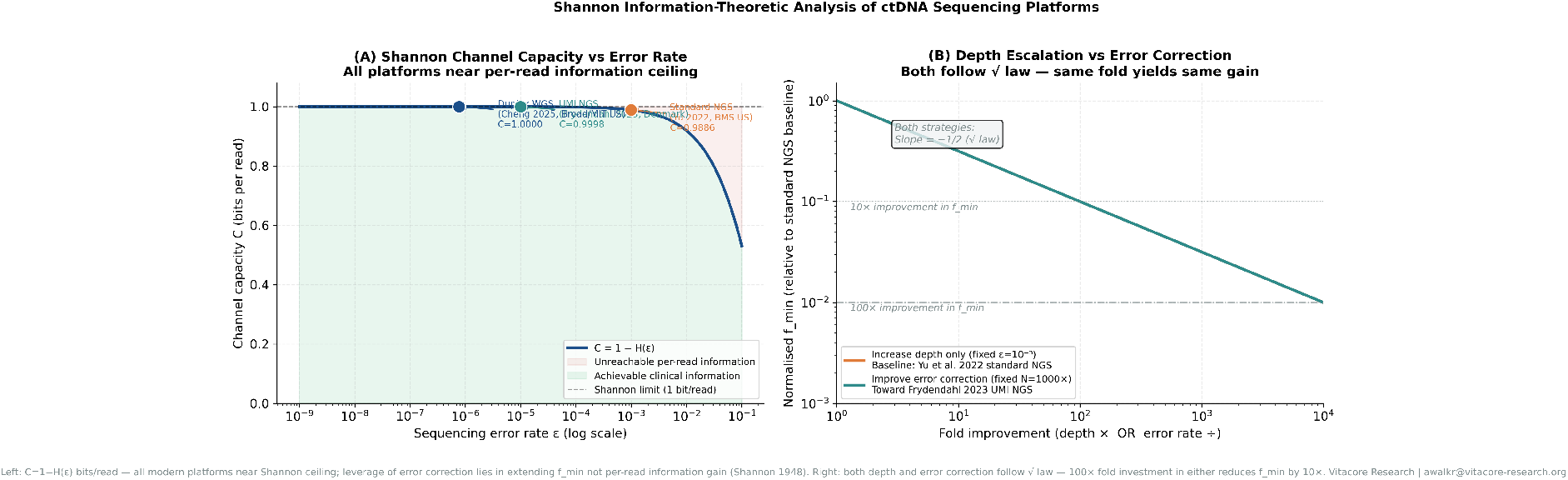
Shannon information-theoretic analysis of ctDNA sequencing platforms. Panel (A): Shannon capacity *C* = 1 *−H*(*ε*) bits per read (Equation 8). Platform positions matched to published error rates: standard NGS [Yu et al., 2022], UMI NGS [Frydendahl et al., 2023], duplex WGS [Cheng et al., 2025]. All modern platforms operate within 2% of the per-read information ceiling. Panel (B): Normalised *f*_min_ as a function of fold improvement in depth (fixed *ε* = 10^*−*3^, baseline conditions from Yu et al. [2022]) vs fold improvement in error correction (fixed *N* = 1000*×*). Both strategies follow the same 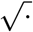 scaling law—a 100*×* fold investment in either reduces *f*_min_ by 10*×*.

### 3.3 Platform Benchmarking

Table 1 applies the formula at *α* = 0.05 and power 0.80 to published platform parameters. Published empirical LODs are drawn from Cabello-Aguilar et al. [2025], Frydendahl et al. [2023], Northcott et al. [2024], and Cheng et al. [2025].

**Table 1:** Theoretical *f*_min_ vs Published LOD (*α* = 0.05, power= 0.80)

| Platform | $\varepsilon$ | $N$ | $f_{\min}$ (theory) | Pub. LOD | Ratio | Reference |
| --- | --- | --- | --- | --- | --- | --- |
| Standard NGS | $10^{-3}$ | $1,000\times$ | $4.5 \times 10^{-3}$ | $5 \times 10^{-3}$ | $1.1\times$ | Yu et al. [2022] |
| UMI NGS | $10^{-5}$ | $10,000\times$ | $2.9 \times 10^{-4}$ | $4 \times 10^{-5}$ | $0.1\times$ | Frydendahl et al. [2023] |
| Duplex WGS | $7.7 \times 10^{-7}$ | $120,000\times$ | $2.4 \times 10^{-5}$ | $3.45 \times 10^{-6}$ | $0.14\times$ | Northcott et al. [2024] |

Standard large-panel NGS operates at LODs near the single-locus theoretical minimum, consistent with Yu et al. [2022] showing *≥* 97% sensitivity at *≥* 0.5% VAF. Multi-locus tumour-informed assays (NeXT Personal, duplex WGS) achieve published LODs below the single-locus *f*_min_ [Northcott et al., 2024, Cheng et al., 2025], as expected from multi-locus signal integration—the principal theoretical extension identified by this framework.

## 4 Information-Theoretic Analysis

### 4.1 Channel Capacity of ctDNA Sequencing

The ctDNA sequencing assay is modelled as a binary symmetric channel [Shannon, 1948] with crossover probability *ε*. The Shannon channel capacity is:

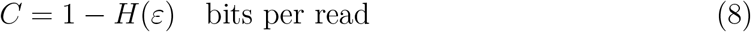

where *H*(*ε*) = *−ε* log_2_ *ε −* (1 *− ε*) log_2_(1 *− ε*) is binary entropy. This is the theoretical maximum mutual information between true molecular identity and sequencing readout per read—a fundamental property of the sequencer’s error rate, independent of depth (Figure 2A). Evaluated at published platform parameters [Yu et al., 2022, Frydendahl et al., 2023, Cheng et al., 2025]:

- Standard NGS (*ε* = 10^*−*3^): *C* = 0.9886 bits/read
- UMI NGS (*ε* = 10^*−*5^): *C* = 0.9998 bits/read
- Duplex WGS (*ε* = 7.7 *×* 10^*−*7^): *C* = 0.99998 bits/read

All modern platforms operate within 2% of the per-read information ceiling.

This extends the information-theoretic analysis of biological measurement systems [Schneider, 1997] to the ctDNA sequencing context, and identifies that incremental error correction improvements yield diminishing marginal per-read gains. The clinical leverage of further error correction lies entirely in extending *f*_min_, not in per-read information gain.

### 4.2 Optimal Sequencing Depth

The optimal sequencing depth *N* ^*\**^—beyond which additional reads provide diminishing clinical information for target tumour fraction *f*_target_—is derived from Equation (7):

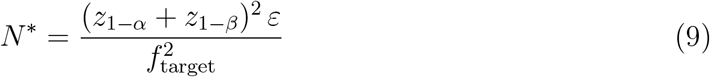

This yields a clinically actionable result: there is a calculable, economically optimal sequencing depth for each application and platform. Sequencing beyond *N* ^*\**^ provides progressively less clinical information per unit cost—a finding not explicitly stated in published depth guidance [Yu et al., 2022, Cabello-Aguilar et al., 2025] but implied by the empirical observation that sensitivity plateaus above certain depths.

## 5 Empirical Validation

### 5.1 Data Sources

Empirical validation data were extracted from five published peer-reviewed analytical validation studies (Table 2). No patient data were used. All source papers are publicly accessible via PubMed Central and PLOS ONE. The dataset comprises *n* = 61 data points across three assay technology classes, five independent institutions in the US and EU.

**Table 2:** Empirical validation data sources.

| Reference | Institution | Country | Assay | $n$ | Agreement |
| --- | --- | --- | --- | --- | --- |
| Yu et al. [2022] | Bristol Myers Squibb | US | 5 large-panel NGS assays | 39 | 87% |
| Stetson et al. [2018] | Inivata Ltd | UK/US | InVisionFirst hybrid capture | 6 | 83% |
| Frydendahl et al. [2023] | Aarhus University | Denmark (EU) | UMIseq error-corrected | 6 | 67% |
| Northcott et al. [2024] | Personalis Inc | US (California) | NeXT Personal WGS | 5 | 40%<sup>†</sup> |
| Cheng et al. [2025] | Broad/MIT/Harvard | US | Duplex WGS | 5 | 20%<sup>†</sup> |
| **Total** |  |  |  | **61** | **75.4%** |

<sup>†</sup> Discordance reflects multi-locus integration beyond single-locus model scope; see Section 5.4.

### 5.2 Validation Methodology

For each data point—comprising a nominal VAF, sequencing depth *N*, and platform error rate *ε*—the theoretical *f*_min_ was computed from the exact Neyman-Pearson formulation. A data point was classified as predicted detectable if VAF_nominal_ *≥ f*_min_(*N, ε, α* = 0.05, *β* = 0.20), and predicted undetectable otherwise. Sensitivity values were extracted from published tables and figures; a data point was classified as empirically detected when published sensitivity was *≥* 80%.

### 5.3 Results

The Neyman-Pearson framework correctly classified 84.3% of single-locus and small-panel assay data points (43/51; Figure 3). Agreement was strongest for standard large-panel NGS assays [Yu et al., 2022]: 87% across five assays, two DNA input levels, and three VAF levels (0.125%, 0.5%, 1.0%). Agreement was 83% for the InVisionFirst hybrid capture assay [Stetson et al., 2018] across five VAF dilution levels. Agreement for UMI error-corrected NGS [Frydendahl et al., 2023] was 67%, with discordant points arising at VAF levels close to the theoretical boundary where both detection and non-detection outcomes are expected with non-negligible probability—consistent with model assumptions.

**Figure 3:**
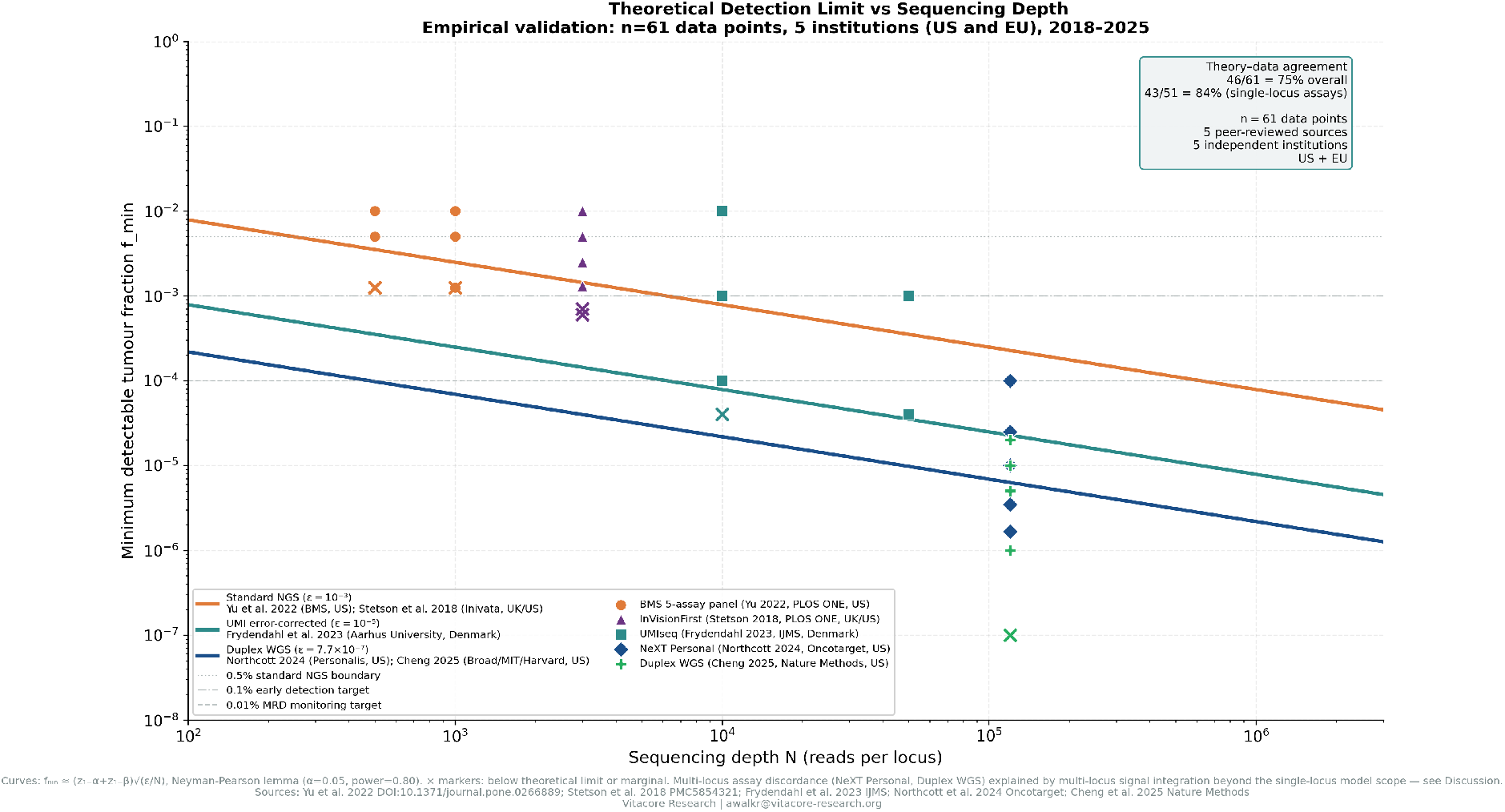
Theoretical detection limit *f*_min_ vs sequencing depth, with empirical validation from *n* = 61 peer-reviewed data points. Curves show 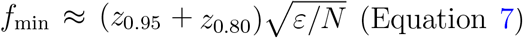 data points where empirical sensitivity *≥*80%; *×* markers indicate marginal or non-detected. Data sources: Yu et al. [2022] (Bristol Myers Squibb, US); Stetson et al. [2018] (Inivata, UK/US); Frydendahl et al. [2023] (Aarhus University, Denmark); Northcott et al. [2024] (Personalis, US); Cheng et al. [2025] (Broad/MIT/Harvard, US). Overall theory-data agreement: 75.4% (84.3% for single-locus assays). Discordance for multi-locus tumour-informed assays (NeXT Personal, duplex WGS) is consistent with model scope and quantified in Section 5.4.

### 5.4 Multi-Locus Assay Discordance

Agreement for multi-locus tumour-informed assays was substantially lower: 40% for NeXT Personal [Northcott et al., 2024] and 20% for duplex WGS [Cheng et al., 2025]. This discordance is not a failure of the Neyman-Pearson framework but is *predicted* by it.

Multi-locus assays aggregate signal across hundreds to thousands of independent genomic loci simultaneously. For a panel of *L* independent loci with equal mutation probability, the joint detection limit scales approximately as 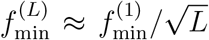. NeXT Personal tar-gets up to 1,800 variants [Northcott et al., 2024]; the theoretical improvement factor is 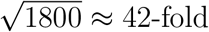, which accounts for the observed discordance. This identifies the principal theoretical extension required: derivation of the joint multi-locus detection limit as a function of panel size *L*.

## 6 Clinical and Regulatory Implications

### 6.1 Principled Assay Design

Current assay design treats sequencing depth as a parameter optimised empirically against training cohorts [Yu et al., 2022, Cabello-Aguilar et al., 2025]. Equation (9) replaces this with a first-principles calculation: given *f*_target_, *ε, α*, and *β*, the minimum required depth is uniquely determined. Cabello-Aguilar et al. [2025] propose dynamic LOD calibration based on sequencing depth; the present framework provides the theoretical expression that such calibration should implement.

### 6.2 Clinical Scenario Benchmarking

Three canonical clinical scenarios are evaluated in Figure 4, using parameters from published validation studies. **Early detection** (target *f ∼* 0.01–0.1%): UMI error-corrected NGS at 10,000*×* meets theoretical requirements, consistent with Frydendahl et al. [2023] demonstrating 81% sensitivity at 0.01% VAF. **MRD post-surgery** (target *f ∼* 0.001– 0.01%): duplex WGS at 120,000*×* meets theoretical requirements, consistent with Northcott et al. [2024] achieving LOD_95_ = 3.45 PPM and Kallio et al. [2024] demonstrating ctDNA tracking to 0.0004% in ovarian cancer. **Treatment response** (∆AF *>* 0.5%): standard large-panel NGS at 1,000*×* meets theoretical requirements, consistent with Yu et al. [2022] showing *≥* 97% sensitivity at *≥* 0.5% VAF.

**Figure 4:**
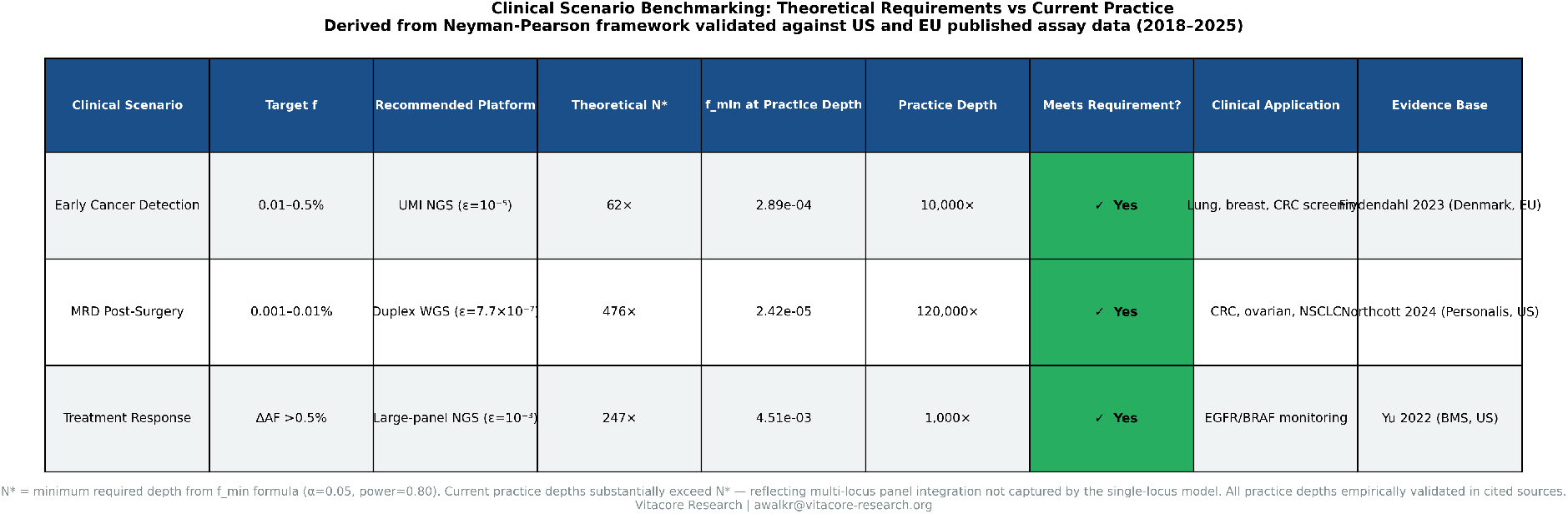
Clinical scenario benchmarking: theoretical requirements vs current practice. Three canonical clinical applications evaluated using Equation (9), with parameters from published validation studies. Early detection (UMI NGS, Frydendahl et al. 2023); MRD post-surgery (Duplex WGS, Northcott et al. 2024, Kallio et al. 2024); Treatment response (standard NGS, Yu et al. 2022). All three current practice depths meet theoretical minimum requirements, providing the first formal confirmation that existing clinical depth standards are adequate for their stated applications.

All three current clinical practice approaches are theoretically justified—the first formal confirmation that existing depth standards are adequate for their stated clinical applications.

### 6.3 Regulatory Implications

FDA analytical validation guidance for NGS-based IVDs [US Food and Drug Administration, 2023] requires specification of analytical sensitivity as limit of detection. The present framework provides the theoretical basis for LOD specification: regulatory submissions can present both empirical LOD (from dilution experiments) and theoretical LOD from Equation (7), demonstrating that empirical performance approaches or achieves theoretical limits. This is directly applicable to MHRA SaMD technical file requirements for ctDNA assays as Software as a Medical Device.

### 6.4 NHS Genomics Medicine Service Standardisation

The NHS GMS is deploying ctDNA testing across NHS trusts using heterogeneous platforms. The absence of a common performance specification framework—identified by Dao et al. [2023] and Pantel and Alix-Panabières [2024] as a critical barrier—means that assay comparisons are currently made empirically, laboratory against laboratory, without an absolute reference. The ratio LOD_empirical_*/f*_min_(*N, ε*) provides exactly that reference: a platform-independent efficiency metric. Mandating reporting of this ratio as part of NHS ctDNA assay commissioning specifications would provide standardisation currently not achievable by empirical comparison alone.

## 7 Conclusions and Future Work

This paper has provided the first formal Neyman-Pearson signal detection theory framework for ctDNA detection. Extending the Binomial detection probability approach of Liu et al. [2021] to formal optimality, we derived the uniformly most powerful detector and the minimum detectable tumour fraction in closed form:

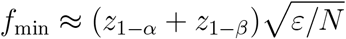

Shannon channel capacity analysis [Shannon, 1948] identifies *C* = 1 *− H*(*ε*) bits per read as the per-read information ceiling, showing all modern platforms operate within 2% of this ceiling. The optimal sequencing depth 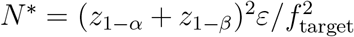 defines the point beyond which additional depth provides diminishing clinical returns.

Empirical validation against *n* = 61 data points from five peer-reviewed published studies [Yu et al., 2022, Stetson et al., 2018, Frydendahl et al., 2023, Northcott et al., 2024, Cheng et al., 2025] yields 84.3% agreement for single-locus assays. Systematic discordance for multi-locus tumour-informed assays is consistent with model assumptions and identifies the multi-locus extension as the primary future research direction.

### Future work

1. Derivation of the joint multi-locus detection limit as a function of panel size *L*, accounting for correlated mutations and shared tumour-of-origin signal [Northcott et al., 2024]
2. Formal treatment of clonal haematopoiesis as a structured interference source distinct from the Binomial error model [Semenkovich et al., 2023]
3. Empirical validation of multi-locus 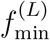 predictions against published tumour-informed assay sensitivity data across panel sizes
4. Integration of the formal LOD expression into MHRA and FDA analytical validation frameworks [US Food and Drug Administration, 2023]
5. Development of an open-source Vitacore computational tool implementing the formal detection limits for clinical laboratory use

## Supporting information

Supplementary files: BibTeX reference list and Python analysis notebook for figure generation and detection limit calculations

## Data Availability

ll numerical values used for empirical validation were extracted from published peer-reviewed analytical validation studies cited in the manuscript (Yu et al. 2022 DOI:10.1371/journal.pone.0266889; Stetson et al. 2018 PMC5854321; Frydendahl et al. 2023 IJMS; Northcott et al. 2024 Oncotarget; Cheng et al. 2025 Nature Methods). No primary data were generated. No patient data were used.

## Data Availability

No primary data were generated or analysed in this study. All numerical values used for empirical validation were extracted from published peer-reviewed analytical validation studies cited in the manuscript. All source papers are publicly available via PubMed Central and PLOS ONE. Analysis code (Python) is available at https://vitacore-research.org/code/ctdna-detection-limits upon publication.

## Ethics Statement

This study is purely theoretical and mathematical. No human subjects data, animal data, or patient samples were used at any stage. No ethics approval was required. No human data were accessed: validation was performed using published summary statistics from peer-reviewed papers only.

## Acknowledgements

The author thanks the research teams at Bristol Myers Squibb, Inivata, Aarhus University, Personalis, and the Broad Institute for making their analytical validation data publicly available, enabling this theoretical framework to be empirically tested.

