## Supplementary files: BibTeX reference list and Python analysis notebook for figure generation and detection limit calculations for "Formalising Limits of Circulating Tumour DNA Detection: A Signal Detection Framework for Clinical Threshold Specification": references.bib

% ── CORE SIGNAL PROCESSING THEORY ─────────────────────────────────────────────

@book{kay1998,
  author    = {Kay, Steven M.},
  title     = {Fundamentals of Statistical Signal Processing, Volume {II}: Detection Theory},
  publisher = {Prentice Hall},
  address   = {Upper Saddle River, NJ},
  year      = {1998},
  isbn      = {978-0135041352}
}

@article{shannon1948,
  author  = {Shannon, Claude E.},
  title   = {A Mathematical Theory of Communication},
  journal = {Bell System Technical Journal},
  volume  = {27},
  number  = {3},
  pages   = {379--423},
  year    = {1948},
  doi     = {10.1002/j.1538-7305.1948.tb01338.x}
}

@article{schneider1994,
  author  = {Schneider, Thomas D.},
  title   = {Information Content of Individual Genetic Sequences},
  journal = {Journal of Theoretical Biology},
  volume  = {189},
  number  = {4},
  pages   = {427--441},
  year    = {1997},
  doi     = {10.1006/jtbi.1997.0540}
}

% ── PRIOR WORK ON BINOMIAL DETECTION IN ctDNA ─────────────────────────────────

@article{liu2021,
  author  = {Liu, Zhe and Qiu, Weijin and Fu, Shujin and Zhao, Xia and
             Xia, Jun and Geng, Chunyu and Yu, Youqian and Li, Ziling and
             Li, Mingzhu and Jiang, Hui and Chen, Fang},
  title   = {Influence of Sequencing Depth on the Fidelity and Sensitivity
             of 1\%--5\% Low-Frequency Mutation Detection and Recommendation
             for Standardization of Sequencing Depth},
  journal = {bioRxiv},
  year    = {2021},
  doi     = {10.1101/2021.12.26.474219},
  note    = {Key prior work: Binomial computation of detection probability.
             This paper extends to Neyman-Pearson optimality and closed-form
             f-min derivation. VERIFY: check final publication status.}
}

% ── EMPIRICAL VALIDATION DATASETS ─────────────────────────────────────────────

@article{yu2022,
  author  = {Yu, Lizhi and Lopez, Gonzalo and Rassa, John and Wang, Yixin and
             Basavanhally, Tara and Browne, Andrew and Huang, Chang-Pin and
             Dorsey, Lauren and Jen, Jin and Hersey, Sarah},
  title   = {Direct Comparison of Circulating Tumor {DNA} Sequencing Assays
             with Targeted Large Gene Panels},
  journal = {PLOS ONE},
  volume  = {17},
  number  = {4},
  pages   = {e0266889},
  year    = {2022},
  doi     = {10.1371/journal.pone.0266889},
  note    = {Bristol Myers Squibb, US. 5 assays, 3 VAF levels. Primary
             validation dataset. VERIFY: confirm full author list.}
}

@article{stetson2018,
  author  = {Stetson, Daniel and Ahmed, Ahad and Xu, Xiaowei and Nuttall,
             Brian R. B. and Lubinski, Thomas J. and Johnson, James H. and
             Barrett, J. Carl and Emery, Stephen C. and Veltri, Robert W. and
             Bhatt, Devashish L.},
  title   = {Analytical Validation of a Next Generation Sequencing Liquid
             Biopsy Assay for High Sensitivity Broad Molecular Profiling},
  journal = {PLOS ONE},
  volume  = {13},
  number  = {3},
  pages   = {e0193802},
  year    = {2018},
  doi     = {10.1371/journal.pone.0193802},
  note    = {Inivata Ltd, Cambridge UK and NC US. InVisionFirst assay.
             VERIFY: check full author list against PMC5854321.}
}

@article{frydendahl2023,
  author  = {Frydendahl, Amanda and Kronborg, Camilla and
             S{\o}rensen, Boe Sandahl and Bech, Boe Steen and
             {\O}sterlind, Kell and Pallisgaard, Niels},
  title   = {Error-Corrected Deep Targeted Sequencing of Circulating
             Cell-Free {DNA} from Colorectal Cancer Patients for Sensitive
             Detection of Circulating Tumor {DNA}},
  journal = {International Journal of Molecular Sciences},
  volume  = {24},
  number  = {4},
  pages   = {3888},
  year    = {2023},
  doi     = {10.3390/ijms24043888},
  note    = {Aarhus University, Denmark. UMIseq error-corrected NGS.
             VERIFY: confirm full author list and page numbers.}
}

@article{northcott2024,
  author  = {Northcott, Josette M. and Bartha, Gabor and Harris, Jason and
             Li, Conan and Navarro, Fabio C. P. and Pyke, Rachel Marty and
             Hong, Manqing and Zhang, Qi and Ma, Shuyuan and Chen, Tina X. and
             Lai, Janet and Udar, Nitin and Saldivar, Juan-Sebastian and
             Ayash, Erin and Anderson, Joshua and Li, Jiang and Cui, Tiange and
             Le, Tu and Chow, Ruthie and Velasco, Randy Jerel and
             Santiago, Rose and Bruce, Robert C. and Goodman, Laurie J. and
             Chen, Yi and Norton, Dan and Chen, Richard O. and Lyle, John M.},
  title   = {Analytical Validation of {NeXT Personal},
             an Ultra-Sensitive Personalized Circulating Tumor {DNA} Assay},
  journal = {Oncotarget},
  volume  = {15},
  pages   = {200--218},
  year    = {2024},
  doi     = {10.18632/oncotarget.28565},
  note    = {Personalis Inc, California US. LOD 1.67 PPM, LOD95 3.45 PPM.
             PMC10939476.}
}

@article{cheng2025,
  author  = {Cheng, Alexandre P. and others},
  title   = {Error-Corrected Flow-Based Sequencing at Whole-Genome Scale
             and its Application to Circulating Cell-Free {DNA} Profiling},
  journal = {Nature Methods},
  year    = {2025},
  doi     = {10.1038/s41592-024-02576-2},
  note    = {Broad Institute, MIT, Harvard, US. Duplex error rate 7.7e-7.
             VERIFY: confirm DOI and full author list.}
}

% ── CLINICAL REVIEWS AND CONTEXT ──────────────────────────────────────────────

@article{pantel2024,
  author  = {Pantel, Klaus and Alix-Panabieres, Catherine},
  title   = {Minimal Residual Disease as a Target for Liquid Biopsy in
             Patients with Solid Tumours},
  journal = {Nature Reviews Clinical Oncology},
  volume  = {21},
  pages   = {175--187},
  year    = {2024},
  doi     = {10.1038/s41571-023-00847-9},
  note    = {VERIFY: confirm page numbers and DOI.}
}

@article{dao2023,
  author  = {Dao, Jonathan and Finlayson, Neil and Bagnato, Dana and
             Gilad-Opher, Anat and Mayer, Alexandra and Sturtevant, Elisa and
             Peled, Nir},
  title   = {Using {cfDNA} and {ctDNA} as Oncologic Markers: A Path to
             Clinical Validation},
  journal = {International Journal of Molecular Sciences},
  volume  = {24},
  number  = {11},
  pages   = {9368},
  year    = {2023},
  doi     = {10.3390/ijms24119368},
  note    = {VERIFY: confirm author list against published version.}
}

@article{cabello2025,
  author  = {Cabello-Aguilar, Simon and others},
  title   = {Real-World Technical Hurdles of {ctDNA NGS} Analysis:
             Lessons from Clinical Implementation},
  journal = {Diseases},
  year    = {2025},
  note    = {VERIFY: confirm journal, volume, and full author list.}
}

@article{sato2025,
  author  = {Sato, Yoshiharu},
  title   = {Liquid Biopsy for Minimal Residual Disease and Monitoring
             in Early-Stage Non-Small Cell Lung Cancer},
  journal = {Exploration of Medicine},
  year    = {2025},
  note    = {VERIFY: confirm journal name, volume, and issue.}
}

@article{slater2024,
  author  = {Slater, Sarah and others},
  title   = {Tissue-Free Liquid Biopsies for Minimal Residual Disease
             Detection in Early Colorectal Cancer: {TRACC} Part {B}},
  journal = {Clinical Cancer Research},
  year    = {2024},
  note    = {VERIFY: confirm full author list, volume, and DOI. n=214 CRC.}
}

@article{wan2020,
  author  = {Wan, Jonathan C. M. and Heider, Katrin and Gale, Davina and
             Murphy, Sean and Fisher, Edward and Mouliere, Florent and
             Ruiz-Valdepenas, Andrea and Moore, Elizabeth H. and Morris,
             James and Chandrananda, Dineika and Rosenfeld, Nitzan},
  title   = {{ctDNA} Monitoring Using Patient-Specific Sequencing and
             Integration of Variant Reads},
  journal = {Science Translational Medicine},
  volume  = {12},
  number  = {553},
  pages   = {eaaz8084},
  year    = {2020},
  doi     = {10.1126/scitranslmed.aaz8084}
}

@article{nordentoft2024,
  author  = {Nordentoft, Iver and others},
  title   = {Whole-Genome Mutational Analysis for Tumor-Informed Detection
             of Circulating Tumor {DNA} in Patients with Urothelial Carcinoma},
  journal = {European Urology},
  year    = {2024},
  note    = {VERIFY: confirm full author list, volume, and DOI.
             91 percent sensitivity, 131-day lead time.}
}

@article{kallio2024,
  author  = {Kallio, Heidi and others},
  title   = {Sensitive Circulating Tumor {DNA}-Based Residual Disease
             Detection in Epithelial Ovarian Cancer},
  journal = {Life Science Alliance},
  year    = {2024},
  note    = {VERIFY: confirm full author list and DOI.
             ctDNA tracked to 0.0004 percent.}
}

@article{semenkovich2023,
  author  = {Semenkovich, Nicole P. and others},
  title   = {Genomic Approaches to Cancer and Minimal Residual Disease
             Detection Using Circulating Tumor {DNA}},
  journal = {Journal for Immunotherapy of Cancer},
  year    = {2023},
  note    = {VERIFY: confirm full author list, volume, and DOI.}
}

@article{tebar2023,
  author  = {T{\'e}bar-Mart{\'i}nez, Roberto and others},
  title   = {Strategies for Improving Detection of Circulating Tumor {DNA}
             Using Next Generation Sequencing},
  journal = {Cancer Treatment Reviews},
  year    = {2023},
  note    = {VERIFY: confirm full author list, volume, and DOI.}
}

% ── REGULATORY ────────────────────────────────────────────────────────────────

@techreport{fda2023,
  author      = {{US Food and Drug Administration}},
  title       = {Considerations for the Design, Development, and Analytical
                 Validation of Next Generation Sequencing-Based In Vitro
                 Diagnostics},
  institution = {US Food and Drug Administration},
  year        = {2023}
}
